# Prevalence of antibodies against sars-cov-2 in professionals of a public health laboratory at são paulo, sp, brazil

**DOI:** 10.1101/2020.10.19.20213421

**Authors:** Valéria Oliveira Silva, Elaine Lopes de Oliveira, Marcia Jorge Castejon, Rosemeire Yamashiro, Cintia Mayumi Ahagon, Giselle Ibette López-Lopes, Edilene Peres Real da Silveira, Marisa Ailin Hong, Maria do Carmo Timenetsky, Carmem Aparecida Freitas, Luís Fernando de Macedo Brígido

**Affiliations:** Adolfo Lutz Central Institute, São Paulo, Brazil: Center of Virology; Center of Imunology

**Author notes:** Corresponding author: Luís Fernando de Macedo Brígido M.D. Ph. D., Adolfo Lutz Institute - Virology Center Ave. Dr. Arnaldo 355, São Paulo, São Paulo, Brazil 05411001 email. All authors approved the final version of the paper. Funding sources: No external funding was available for this work. Ethical Approval: IAL Ethical Committee CAAE : 31924420.8.0000.0059.

**Keywords:** COVID-19, SARS-CoV-2, Seroprevalence, Survey, Diagnosis, antibodies

## Abstract

**Background:** Covid-19 Serology may document exposure and perhaps protection to the virus and serological test may help understand epidemic dynamics. We tested health workers form a public laboratory to evaluate previous exposure to the virus and estimate the prevalence of antibodies against-SARS-CoV-2 in Adolfo Lutz Institute, State of São Paulo, Brazil.

**Methods:** This study was an open, prospective evaluation among professionals of Adolfo Lutz Institute some administrative personnel from the Secretary of Health that shares common areas with the institute. We used a lateral flow immunoassay (rapid test) to detect IgG and IgM for SARS-CoV-2; positive samples were further evaluated using Roche Electrochemiluminescence assay. SARS-CoV-2 RNA by real time reverse transcriptase polymerase chain reaction (RT-PCR) was also offered to participants.

**Results:** A total of 406 HPs participated. Thirty five (8.6%) tested positive on rapid test and 32 these rapid test seropositive cases were confirmed by ECLIA. 43 HPs had SARS-CoV-2 RNA detected at a median of 33 days, and the three cases not reactive at Roche ECLIA had a previous positive RNA. Outsourced professionals (34% seropositive), males (15%) workers referring COVID-19 patients at home (22%) and those living farther form the institute tended to have higher prevalence of seropositivity, but in multivariable logistic analysis only outsourced workers and those with COVID patients at home remained independently associated to seropositivity. We observed no relation of seropositivity to COVID samples handling. Presence of at least one symptom was common but some clinical manifestations as anosmia/dysgeusia. Fatigue, cough and fever were associated to seropositivity.

**Conclusions:** We documented a relatively high (8.6%) of anti-SARS-CoV-2 serological reactivity in this population, higher among outsourced workers and those residing with COVID-19 patients. COVID related work did not increased seropositivity. Some symptoms show strong association to COVID-19 serology and may be used in scoring tools for screening or diagnosis in resort limited settings.

## 1. Introduction

Over one million deaths due to the COVID-19 pandemic have been reported in by the end of September 2020 (OMS, 2020). Severe Acute Respiratory Syndrome Coronavirus 2 (SARS-CoV-2), the cause of COVID-19, has spread across Brazil and since the first recorded case in February 26, 2020 (Brasil^1^, 2020). Contrary to places as China and parts of Europe, Brazil never achieved a true decline in new cases incidence, and the country has plateaued at a high rate of about 32.058 new cases every day (OMS, 2020). In Brazil, there have been over four millions documented cases of COVID-19 and more than 140 thousand deaths at the end of September 2020 (Brasil^2^, 2020) across all its territory, but numbers of cases are probably underestimated due to testing limitations.

The search for markers of immunity and diagnosis with serology has led to development of different assays, but specificity and sensibility issues have been reported (Castro et al., 2020). Along with the unknown nature of a protective immunity and the fact that antibodies emerge at the end, or after, infectiousness phase of the disease, has hamper the use of serology as a diagnostic clinical tool. However, it remained valid to evaluate population exposure to the virus, guiding public health policies and may provide a general framework for understanding virus exposure.at a population level. The performance of systematic and comprehensive tests to identify the infection in health professionals (HP) and other key areas is important, even if they are not in direct contact with patients, and available to estimate the prevalence of infected and virus transmission in within health services. A meta-analysis of eleven studies showed that almost 10% of COVID-19 positive patients are health professionals (Sahu et al., 2020).

The Adolfo Lutz Institute, the Central laboratory of public health of State of São Paulo, has increased its activities to fight COVID since early in the pandemic. Nowadays, the institute and it regional centers receives most respiratory tract samples collected in the São Paulo State, processes part of the daily load and distribute the remaining to associated clinical laboratories. São Paulo is the most populous State and accordantly has the largest number of COVID-19 cases in the country, with 985.628 documented cases and 35.622 thousand deaths by the end of September 2020 (SEADE, 2020). COVID-19 epidemic in Brazil spread quickly among health workers (Valente et al., 2020; Faíco-Filho et al., 2020) and the increase in infection rates among those professionals has the potential for compromising the health system (Barranco et al., 2020). There is limited data on how the professionals of public health diagnostic and research services / institutes, an example of an active sector during the epidemic, in areas both linked or not to COVID-19 laboratory work. We evaluated the presence of antibodies against-SARS-CoV-2 among professionals of Adolfo Lutz Institute, State of São Paulo, Brazil.

## 2. Materials and Methods

### 2.1. Study population

This study was performed among professionals of Adolfo Lutz Institute in São Paulo, Brazil and administrative personnel of the Secretary of Health that shares common work buildings.

### 2.2. Laboratory tests of SARS-CoV-2 infection

#### 2.2.1. SARS-CoV-2 antibodies

During the period from June 5, 2020 to July 31, 2020, workers we invited to participate in this voluntary survey answered a brief questionnaire, containing demographic data, work activities, symptoms and exposure to COVID-19. Those agreeing to collect peripheral blood samples to test for the presence of antibodies against-SARS-CoV-2 in serum were included. The blood was collected both with and without out anticoagulant and was centrifuged (2000 g x 15 min). We used a commercial antibody test lateral flow immunoassay (LFIA) method, (SARS-CoV-2 Wondfo, Guangzhou Wondfo Biotech Co., Ltd., China) to perform the immunochromatographic assay following the manufacturer’s instructions. This test detects IgG and IgM isotypes that are specific for the SARS-CoV-2 receptor binding domain (RBD). Reagent samples at LFIA were further evaluated using Electrochemiluminescence assay (ECLIA) Elecys Anti-SARS CoV-2 (Roche Diagnostics, Rotkreuz, Switzerland) which is an immunoassay for the in vitro qualitative detection of antibodies (as IgA, IgM and IgG isotypes) that uses a recombinant protein representing the nucleocapsid (N) antigen in a double-antigen sandwich assay format.

#### 2.2.2. SARS-CoV-2 RT-PCR

SARS-CoV-2 RNA by real time reverse transcriptase polymerase chain reaction (RT-PCR) was offered to participants at the time of this survey. Some individuals tested for SARS CoV-2 2 in other laboratories using swab collections and those were also counted as RNA tested in this analysis. RNA was obtained from nasopharyngeal or oropharyngeal secretions either by regular swab collection method (Brasil^3^, 2020), or throat wash (López-Lopes et al., 2020), used for most in-site collections due to swabs sort supply, Briefly, participants received a 5 mL chilled sterile 0.9% saline in a 50 mL falcon tube The contents of the gargle were returned to the tube after approximately 5+ seconds. Participants were instructed to perform the procedure outdoors, at a safe social distance. The tubes were kept at approximately 4-8° C before and after collection, and were processed in the same day. TNA was extracted with a RNA extraction method (Quiagen, USA, Bio Gene, Quibasa, or by automated extraction at Abbott M2000) more recently Quick Extract™ Solution, Lucigen) was used. COVID-19 RNA was retrotranscribed and amplified using the commercial Allplex kit (Seegene, Corea), which is based on the Charité protocol (Corman, et al., 2020). The samples with amplification in the three viral targets (E, RdRP and N) were considered positive. As recommended for the Influenza assay, human RNAse P was used to assess the quality control of the sample and the presence of inhibitors, and human RNAse P cycle thresholds (CTs) up to 37 were considered valid.

### 2.3. Statistical Analysis

We evaluated seroprevalence and its association to demographic and other available information including age, sex, workplace characteristics, contact with confirmed or suspected cases of COVID-19, history of comorbidities (hypertension, diabetes, obesity, autoimmune diseases, among others) and presence of symptoms. Total Number (percentage) or median (interquartile) are shown. Chi-square, Fischer, Kruskal-wallis or Mann Whitney tests, as appropriate. For evaluating the independence of the Association we performed logistic regression with variable showing p value above 0.2 in univariable analysis. The statistical analysis STATA v13.0 program (StataCorp LP, College Station, TX, USA) was used, with a two tailed p <0.05 considered as statistically significant.

### 2.4. Ethical Issue

The study was reviewed and approved by the ethics committee CAAE: 31924420.8.0000.0059 and written informed consent was obtained from all study participants.

## 3.Results

### 3.1. Demographic data among healthcare professionals

A total of 406 individuals participated in the survey. Volunteers were mostly female, 296 (72%) with a median age 50 (IQR 40-57), and 53 (13%) over 60 years old but only 4 over 70. that reflected the composition of institute workers overall. Professionals were classified according to the area of activity as (i) Administrative 82 (20%), (ii) BioMedical laboratories 224 (55%), (iii) Chemistry laboratories 68 (17%) and (iv) 32 (8%) Outsourced workers (including security guards, car valets, cleaning assistants and other support areas). Exposure to individuals symptomatic for COVID-19, or diagnosis was reported by 272 (67%) of the participants.

### 3.2. Antibody response

Thirty five (8.6%) individuals tested positive for (IgM / IgG) serology for SARS-CoV-2. 32 these rapid test seropositive cases were confirmed by ECLIA. Three not reactive at Roche ECLIA had a previous positive RNA, and were considered true positive for the purpose of this analysis. Males, outsourced workers and those referring residing with a symptomatic or diagnosed COVID-19 patient at home tended to have more positive serological results (Figure 1).

**FIGURE 1.**
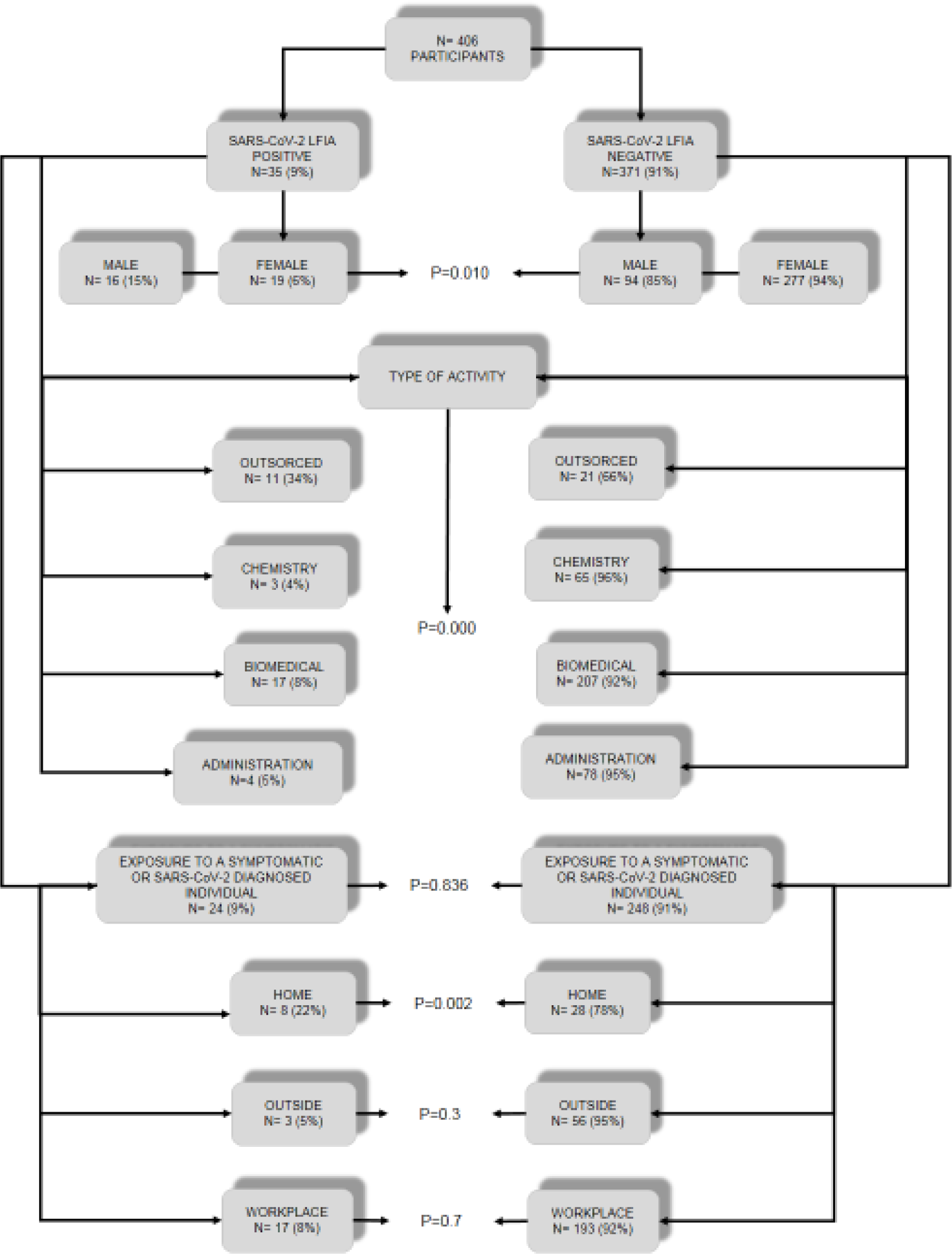
Demographic and COVID cases exposure among study participants according to SARS-CoV-2 antibodies.

### 3.2. Clinical manifestations and presence of SARS-CoV-2 antibodies

One hundred and ninety four (48%) of the interviewees at the time of sample collection reported the presence of at least one symptom. The most frequent were headache in 129 (32%), cough in 67 (17%) and fatigue 70 (17%). Although the presence of a symptoms was more frequent among those tested positive (60%), it was also common among those testing negative (47%, p=0.13). However, some symptoms were associated to seropositivity, as cough, fatigue, fever and especially anosmia/dysgeusia. Table 1 shows the symptoms investigate at questionnaire.

**TABLE 1.**
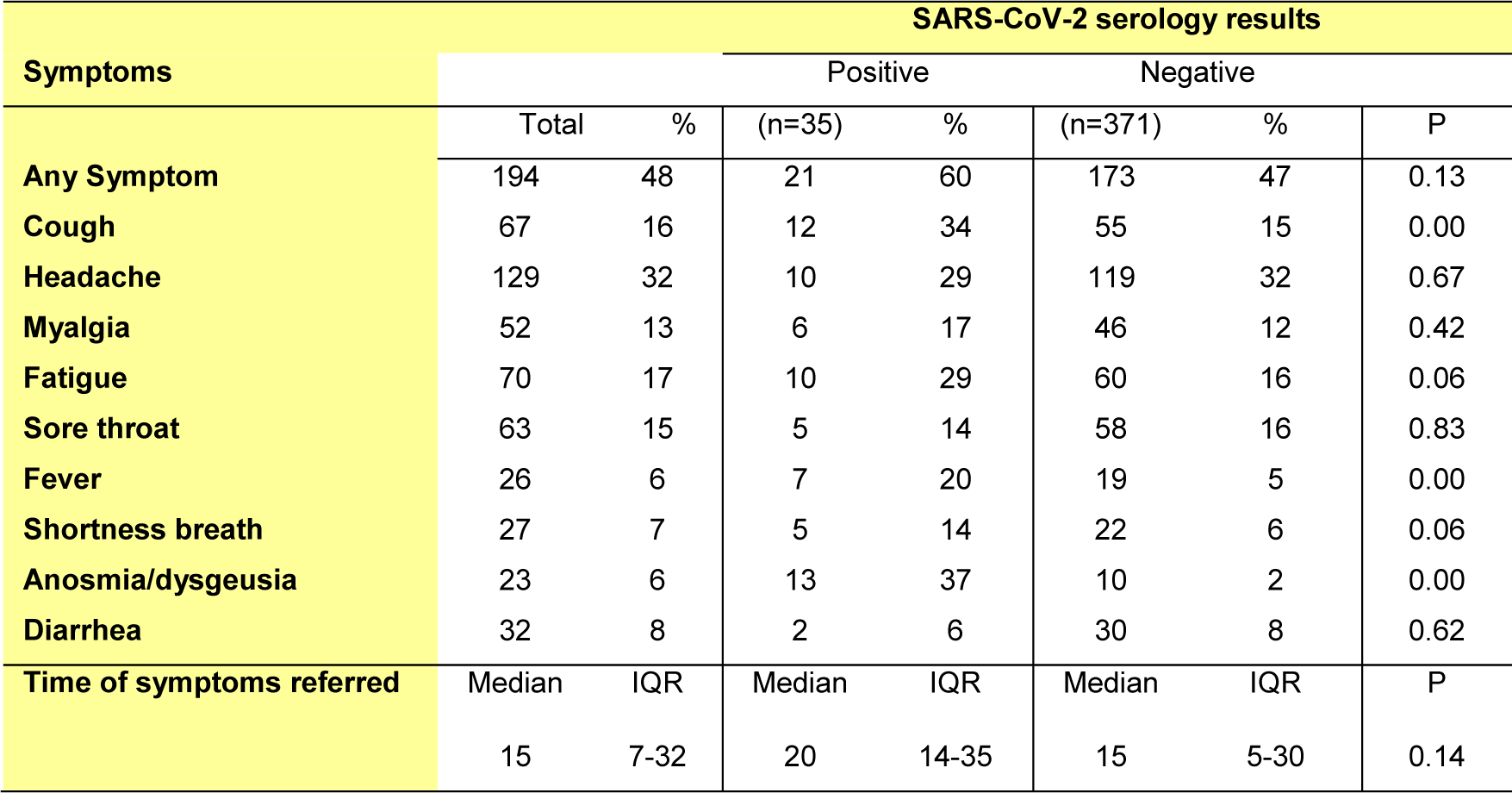
Symptoms presented by individuals, according to serology results of Adolf Lutz Institute professionals.

### 3.3. Correlation between RNA detection results and serology rapid test result

Three hundred and twenty-one participants also had a RNA test collected at the day of serology test or before, with 44 individuals RNA positive. Seropositivity was associated to SARS-CoV-2 RNA detection, with 55% of seropositive cases with previous RNA detection whereas only 4% of those RNA negative were seropositive (p<0.001). Detection of RNA occurred at a median of 33 days (IQR 17-47) before serology.

In table 2 we describe referred clinical symptoms according to serology and RNA results.

**TABLE 2.**
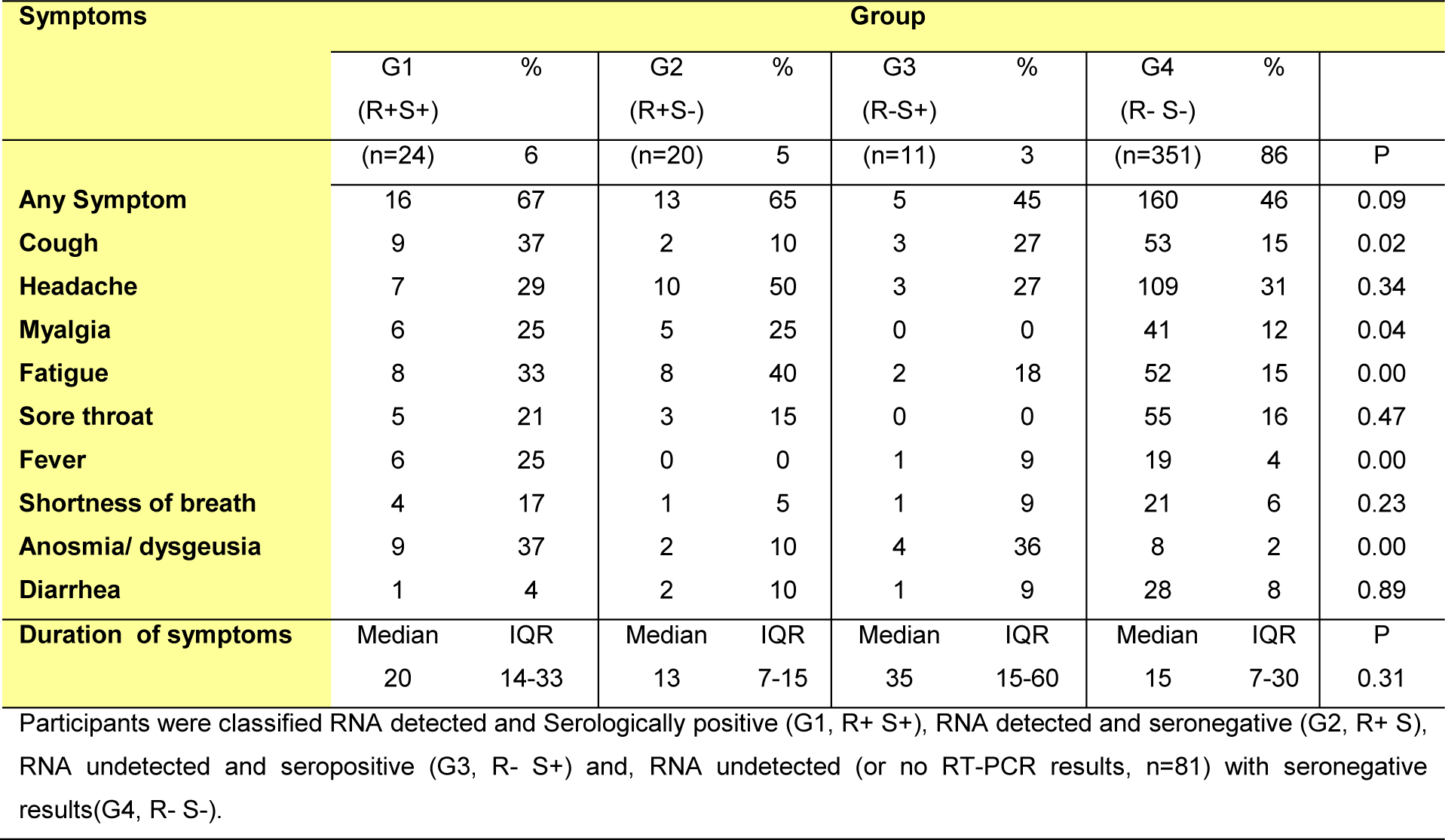
Results of RT-PCR and serology (LFIA) results of Adolfo Lutz Institute professionals.

### 3.4. Correlation between Seropositivity and professional activity

One hundred and forty (34%) of the participants perform activities related to the diagnosis of SARS-CoV-2, of which 102 (73%) perform one or more activity handling biological samples: sample screening (51, 36%), RNA extraction 42 (30%), material preparation 22 (16%), RT-PCR perform 19 (14%), and research 7 (5%). Other activities, which did not involve biological samples, were also mentioned: sample registration and diagnostic report 25 (18%), administrative activities and supply chain management 16 (11%) and, cleaning and decontamination of areas 8 (6%). There was no association COVID-19 serology results and COVID-19 related activity. The differences are not statistically relevant (p=0.122), even when activities were assessed separately: Sample screening (p=0.8), RNA extraction (p=0.3), Material preparation (p=0.4), RT-PCR perform (p=0.6) and research (p=0.4). Considering the handling of *in natura* samples (sample preparation and RNA extraction).

The frequency of comorbidities among participants were: High blood pressure 89 (22%), breathing problems 51 (13%), diabetes 35 (9%), obesity 38 (9%), heart diseases 17 (4%), cancer 4 (1%) and 5 (1%) with immune issues. Fourth-two (10%) individuals reported smoking. Having some comorbidity or smoking was not associated to serology (p=0.88 and p=0.35, respectively).

Three hundred and eighty-eight (97%) disclosed home address and the median distance between the workplace and the home of the interviews was 15 km (IQR 10-20). Considering only those that live in the metropolitan area (circa 50 km from the institute), there was a strong correlation of living far from the institute and doing outsource work (23 km, IQR 16-33) vs (14 Km IQR 10-21) (p=0.0001). Longer distance traveled show some association to seropositivity (p = 0.041). Using public transportation (bus, train and/or subway) to get to the workplace was associated to seropositivity (p=0.3). Table 3 show unadjusted and adjusted logistic analysis of demographic variables associated to COVID-19 seropositivity.

**TABLE 3.** Logistic regression to evaluate the association of demographic variables to seropositivity to SARS-CoV-2.

|  | Demographic factors associated to seropositivity |  |  |  |  |  |
| --- | --- | --- | --- | --- | --- | --- |
|  | Unadjusted |  |  | Adjusted |  |  |
|  | Odds ratio | *p* | 95% CI | Odds ratio | *p* | 95% CI |
| **Outsourced workers** | 7.639 | 0.000 | 3.30 – 17.67 | 6.439 | 0.000 | 2.31 – 17.87 |
| **Longer Distance** | 2.236 | 0.045 | 1.02 – 4.91 | 1.594 | 0.282 | 0.68 – 3.72 |
| **Male Sex** | 2.481 | 0.012 | 1.22 – 5.02 | 1.937 | 0.121 | 0.83 – 4.47 |
| **COVID co-resident** | 3.629 | 0.004 | 1.51 – 8.73 | 4.831 | 0.002 | 1.79 – 13.02 |

Table 3 includes demographic variables with p<0.2 at unavailable, unadjusted analysis as outsourced category vs other workers, shorter vs monger medina distance from work, male vs female sex and reporting residing with a COVID patient vs not, 95% CI for 95% Confidence interval.

## 4. Discussion

The institute is a research and diagnostic reference service of the São Paulo State Health Department kept many working activities during the restriction of non-essential activities, at the initial phase of the COVID-19 pandemic in São Paulo, from March to early June 2020. Although a true lockdown was never attained in this and other metropolitan areas of Brazil, an important decrease in social and economic activities could be noted during this period. The institute, on the other hand, actually increased some of its activities to cope with the fight against the COVID. Now, most of activities in the metropolitan region have been reestablished, albeit with rules for social distance and mask wearing. In this study we documented a seroprevalence of 8.6% in after about 4 month of circulation of SARS-CoV-2 in the region.

Few studies on SARS-CoV-2 seroprevalence are available in Brazil. The larger is a nationwide seroprevalence survey at households, showing much lower of seropositivity, with estimates of 1·9% (95% CI 1·7-2·1) for May to 3·1% (2·8-3·4) for June (Hallal et al., 2020). For the São Paulo area, this study estimated a prevalence from 2% to 4.9% (Hallal et al., 2020). The national estimates included some cities in the amazon basin with prevalence up to 25%, associated to a high case fatality in this period. Another study in southern State, that were not so much affected in this initial phase, showed lower estimates, around of 0.05% to 0.2% (Silveira et al., 2020). Both studies used the same rapid test of our study, but contrary to our study, that used serum, both surveys used blood drops from finger prick that have been documented to underestimate the seroprevalence (Santos et al., 2020).

Surveys that include health workers are more limited in Brazil. In a study with health professionals working on the front line to combat COVID-19 at North of country, the prevalence of antibodies were 21,5% [Melo et al., 2020]. A blood bank detected 4% seropositivity to COVID-19 (Amorim Filho et al., 2020).

A large study in Denmark showed an overall seroprevalence of 3.4% (CI: 2.5%- 3.8%) among health care workers. The rates varied from as high as 29.7% among those with contact with patients to as low as 2.2% in those that not involved in direct contact (Jespersen et al., 2020), which is more close to the characteristics of our study population. Studies in other countries with healthcare workers show prevalence ranging from 1.07 – 17.14% (Lahner et al., 2020; Jeremias et al., 2020; Psichogiou et al., 2020; Pallet et al., 2020; Korth et al., 2020; Chen et al., 2020; Garcia-Basteiro et al., 2020), but laboratory workers were not specifically evaluated.

At the study set up one of the questions was to evaluate if handling biological samples for diagnosis of COVID-19 was related to infection. We evaluate both professional received clinical samples as well as those that perform more risk related activities as preparation of samples for RNA extraction and found no significant association seropositivity. This was also observed in relation to documented infection by rt PCR (López-Lopes et a., 2020).

Previous or current symptoms were commonly reported by either seropositive and negative individuals, but some were associate to seropositivity and can be considered more specific, as cough, fever, fatigue and particularly anosmia and/or dysgeusia. Alterations in smell and taste, although can occur in other pathologies, has been associated as a COVID-19 (Rocke et al., 2020; Russel et al., 2020; Makaronidis et al., 2020) with high prevalence among infected participants (Gómez-Ochoa et al., 2020).

We observed in our study that the presence of symptoms was more reported in those in which RNA was detectable (groups 1 and 2). Cough, myalgia, fatigue, and fever were the most frequent symptoms among participants with detectable RNA. Particularly the presence of cough was also more frequent in-group 4 (negative results in RT-PCR and LFIA) than in group 2, which suggests that presenting this symptom is a criterion for conducting an investigation for COVID-19. Anosmia or/ and dysgeusia and shortness of breath was observed more frequently in the group in which serology was positive (group 1 and 3), suggesting that this symptoms may be correlated with the induction of humoral response (Makaronidis et al., 2020). The median time reported among participants who had anosmia and / or dysgeusia in these groups (1 and 3) was 17 days and 60 days (IQR 8.5-25.5 and IQR 15-90, respectively).

The minority in our study did not report having symptoms before positive serology (14, 40%) and rt-qPCR positive (16, 34%) diagnosis.

Beyond the usefulness of serology to understand the pandemic, the professionals had the opportunity to know their diagnosis, favoring a decrease in the anxiety associated to the pandemic. Moreover, RNA detection allowed swift isolation from colleagues and limited local spread, especially for those asymptomatic.

Transmission is more likely in indoor than outdoor environments (Nishiura et al., 2020) and that home confinement and isolation measures influence due to economic and social aspects, and vary between regions in a country that is heterogeneous like Brazil (Nadanovsky et al., 2020). Our data indicate that living with diagnosed or symptomatic people was more relevant in domestic environment than in the workplace, which can be justified by more intimate contact and extensive sharing of areas, in spite the guidelines advising family members to distance themselves.

Our data point to a similarity with others study about health workers in São Paulo that evaluated positive cases among professionals from different areas of the hospital, regardless of their activities. Thirty-six percent (169/466) of workers who had influenza-like symptoms with suspected COVID-19 tested positive on RT-PCR. Professionals from laboratory areas were 25% while workers who had no direct contact with patients (administrative areas, security and cleaning staff) had a higher infection rate, especially in the areas of logistics (Faíco-Filho, et al., 2020)

In our study, we chose to use a rapid test for preliminary results, despite the its reported performance (Sensitivity: 86, 43% [95% CI: 82, 51%∼89, 58%] and Specificity: 99, 57% [95% CI: 97, 63%∼99,92%]).. We chose to use the serum obtained from the peripheral blood because the test performance was better in terms of sensitivity when used in serum and not in blood capillary, as suggested in others studies (Santos et al., Wu et al., 2020; Serrano et al., 2020). This test may be performed with whole blood, plasma and serum.

We recognize some limitations in our study, as : 1-we used the LFIA method as a preliminary assessment and although confirmed most cases with a second, high throughput ECLIA method, false negative cases would not be identified and we can be under estimating the true prevalence, however, the use of serum instead of finger prick drops may have improved sensibility. 2-Despite our efforts, not all employees joined the study and the prevalence does not reflect the actual SARS-CoV-2 seroprevalence of the institute, but more than half of the workers did contributed to this work. 3-The collection of blood samples was in some cases performed in parallel with the gargle / oropharyngeal smear samples, so we cannot exclude that professionals with a detectable RNA result with a negative LFIA test may have become positive over time. 4-some of the questionnaire were not completed by participants limiting the interpretation of some of the results, but the associations reported were very strong and possibly results were not influenced by missing data.

## 5. Conclusion

We documented a relatively high (8.6%) of anti-SARS-CoV-2 serological reactivity in this population, with higher rates among outsourced workers and those residing with COVID-19 patients. COVID samples handling was not related to increased seropositivity. Some symptoms how strong association to COVID-19 serology and may be used in scoring tools for screening or diagnosis in resort limited settings.

## Data Availability

The data that support the findings of this study are openly available in COVID-19 SARS-CoV-2 preprints from medRxiv and bioRxiv

